# Multiplex PCR amplicon sequencing revealed community transmission of SARS-CoV-2 lineages on the campus of Sichuan University during the outbreak of infection in Chinese Mainland at the end of 2022

**DOI:** 10.1101/2023.05.22.23290366

**Authors:** Langjun Tang, Zhenyu Guo, Xiaoyi Lu, Junqiao Zhao, Yonghong Li, Kun Yang

**Affiliations:** Department of Pharmaceutical & Biological Engineering, School of Chemical Engineering, Sichuan University, Chengdu 610065, China

**Keywords:** Multiplex PCR amplicon sequencing, Wastewater-based epidemiology, SARS-CoV-2, Virus lineages, RT-qPCR

## Abstract

During the pandemic of COVID-19, wastewater-based epidemiology has become a powerful epidemic surveillance tool widely used around the world. However, the development and application of this technology in Chinese Mainland are relatively lagging. Herein, we report the first case of community circulation of SARS-CoV-2 lineages monitored by WBE in Chinese Mainland during the infection outbreak at the end of 2022 after the comprehensive relaxation of epidemic prevention policies. During the peak period of infection, six precious sewage samples were collected from the manhole in the student dormitory area of Wangjiang Campus of Sichuan University. According to the results RT-qPCR, the six sewage samples were all positive for SARS-CoV-2 RNA. Based on multiplex PCR amplicon sequencing, the local transmission of SARS-CoV-2 variants at that time was analyzed. The results show that the main virus lineages in sewage have clear evolutionary genetic correlations. Furthermore, the sampling time is very consistent with the timeline of concern for these virus lineages and consistent with the timeline for uploading the nucleic acid sequences of the corresponding lineages in Sichuan to the database. These results demonstrate the reliability of the sequencing results of SARS-CoV-2 nucleic acid in wastewater. Multiplex PCR amplicon sequencing is by far the most powerful analytical tool of WBE, enabling quantitative monitoring of virus lineage prevalence at the community level.

**Highlights:**

1. Six sewage samples were collected on Wangjiang Campus of Sichuan university at the end of 2022.
2. SARS-CoV-2 nucleic acid was detected in all six sewage samples via qPCR.
3. Multiplex PCR amplicon sequencing reveals the local transmission of SARS-CoV-2 lineages.
4. Multiplex PCR amplicon sequencing is to date the most powerful WBE tool.

**GRAPHICAL ABSTRACT:** 

## 1. Introduction

Since the first detection of SARS-CoV-2 nucleic acid in sewage [1, 2], the wastewater-based epidemiology (WBE) of SARS-CoV-2 has made great progress thanks to the efforts of many scholars around the world [3]. The efforts made in the past two years include but are not limited to the explorations on sampling strategy [4, 5], sample pretreatment [6, 7], nucleic acid extraction [8], nucleic acid detection, decay kinetics of virus (nucleic acid) in sewage [9], correlation between sewage viral RNA concentration and community infection incidence [10] and community transmission of virus lineages [11].

So far, the WBE of SARS-CoV-2 has been developed into four main application scenarios [12, 13]. We summarize them as four “**W**” functions of WBE: to give early warning of the outbreak (**W**hen) [14, 15], to track the trend of community infection (to **W**hat extent) [16], to pinpoint the infection hotspots (**W**here) [17, 18], and to analyze the circulation and evolution of virus lineages (**W**hich variants) in the community [11]. Quantitative tracking the circulation and evolution of virus lineages in communities is currently at the forefront of WBE research, which has fully utilized the qualitative and quantitative capabilities of sewage surveillance. At the beginning, RT-PCR or RT-qPCR targeting specific mutations were used to track a few virus variants of interest [19, 20]. Subsequently, it developed to track mutation sites via high-throughput next-generation sequencing [21-23]. At present, the whole genome sequencing of sewage SARS-CoV-2 can comprehensively track the circulation of different virus lineages in the community [11]. There are currently three whole genome sequencing methods for viruses: multiplex PCR amplicon-based sequencing, hybrid capture-based sequencing, as well as ultra-high-throughput metatranscriptomic sequencing [24]. Multiplex PCR amplicon sequencing, due to its highest enrichment efficiency, is very suitable for whole genome detection of trace microorganisms in complex matrices such as sewage samples. Current sequencing library construction methods include ARTIC [25], Swift [11] and ATOPlex [24]. The sequencing platforms involve MinION, Illumina, ONT-Nanopore and BGI-DNBSEQ.

The development of WBE for SARS-CoV-2 in Chinese Mainland is relatively lagging. There were a few reports about the detection of SARS CoV-2 RNA from hospital wastewater only at the beginning of the epidemic [26-28]. However, in the Hong Kong region, there has been close cooperation between researchers and the government, and a relatively complete sewage virus monitoring network has been formed [29]. The technical methods have also been optimized and are basically becoming mature [6, 8]. Herein, we will report the first detection of community circulation of SARS-CoV-2 lineages via WBE in Chinese Mainland. During the peak period of the nationwide infection outbreak at the end of 2022 after the comprehensive relaxation of epidemic prevention policies, six precious sewage samples were collected in the student dormitory area of Wangjiang Campus of Sichuan University. Through RT-qPCR detection, the six sewage samples were all positive for SARS-CoV-2 nucleic acid. Based on multiplex PCR amplicon sequencing, the local circulation and evolution of SARS-CoV-2 variants was analyzed. To prove the reliability of WBE analysis, we specially tracked the database information about the circulation and evolution of SARS-CoV-2 lineages in Chinese Mainland at that time.

## 2. Materials and methods

### 2.1. Wastewater sampling

From the end of 2022 to the beginning of 2023, Chinese Mainland experienced a peak of SARS-CoV-2 (Omicron) infection. In less than three months, more than 80% of the population was infected [30] (https://doi.org/10.46234/ccdcw2023.070). The main authors of this article were infected during the large-scale infection of SARS-CoV-2 in Chinese Mainland. Langjun Tang developed symptoms on December 9th after being infected and was later quarantined in a school hotel. Considering that the Omicron variant has become much less pathogenic, Chinese government has fully relaxed its epidemic prevention policy throughout that period. Tang is believed to be one of the last COVID-19 patients to be quarantined at the school. He was released from quarantine on December 19th after a negative nucleic acid test. Kun Yang developed symptoms on December 15th, and all serious symptoms basically disappeared on December 18th. Starting from December 27th, untreated wastewater samples were collected every two days from sewage manholes in the dormitory area of Wangjiang campus of Sichuan University, Chengdu, China. Until January 6th, when the school had an early winter vacation, sampling was forced to be terminated due to closed campus management. Tang took full charge of the sample collection. The autosampler was programmed to collect 100 mL wastewater every 1 h over a 24-h period that were composited to provide 24-h composite samples. The sampling instrument has its own refrigeration system, and the sample temperature is maintained at 4 °C during the sampling process. During that period, a total of 6 precious sewage samples were obtained. Collected sewage samples were transported on ice to the laboratory timely and stored frozen at -40 °C for future detection. After subsequent qPCR detection, six sewage samples were all positive for SARS-CoV-2 nucleic acid.

### 2.2. Virus concentration

To recover virus and viral RNA from sewage samples, a modified PEG precipitation method was applied [6]. Typically, the sewage samples were centrifuged at 1680 × g for 5 min to remove large particles. After transferring the supernatant in a new 50-mL centrifuge tube, 3.50 g of PEG 8000 (to a final concentration of 100 g/L) and 0.79 g of NaCl (to a final concentration of 22.5 g/L) were dissolved in 35 ml of supernatant via gentle rotation. Thereafter, virus was precipitated statically overnight at 4 °C. The precipitated virus and viral RNA was recovered in a pellet by centrifugation at 14,000 g for 1 h at 4 °C.

### 2.3. Nucleic acid extraction and qPCR detection

Pellets deposited by centrifugation were extracted nucleic acid using MagicPure^®^ Viral DNA/RNA Kit (TransGen Biotech Co., Ltd, Beijing, China) according to the manufacturer’s instructions. Quantitative polymerase chain reaction (qPCR) was applied to quantify the sewage endogenous biomarker crAssphage using the PerfectStart^®^ Green qPCR SuperMix (TransGen Biotech Co., Ltd, Beijing, China). Reverse transcription qPCR (RT-qPCR) was used to quantify SARS-CoV-2 with the TransScript^®^ II Probe One-Step qRT-PCR SuperMix (TransGen Biotech Co., Ltd, Beijing, China). The primer pairs and probes targeting the genes of SARS-CoV-2 and the crAssphage are listed in Table 1. The protocols for qPCR and RT-qPCR are attached in the supplementary information (Supplementary documents 1 and 2, respectively).

**Table 1.**
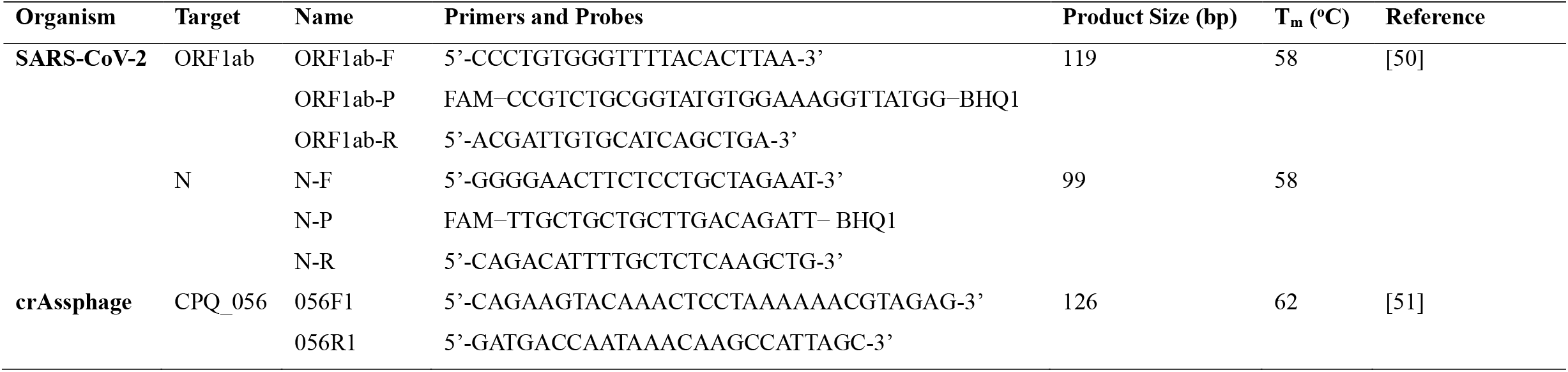
Primer/probe sets targeting the genes of SARS-CoV-2 and crAssphage.

The relative abundance of SARS-CoV-2 in sewage was calculated with the following formula [31].

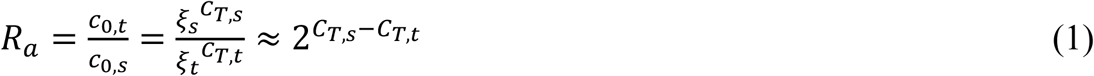

Where *ξ* is the apparent amplification coefficient, which is equal to 2 under ideal conditions; *C*_*T*_ is the threshold cycle number; the subscript *s* indicates the gene of the reference biomarker crAssphage; and the subscript *t* refers to the target virus genes (ORF1ab or N).

### 2.4. ATOPlex multiplex PCR amplicon sequencing

To analyze the whole genome of SARS-CoV-2 in sewage, we applied ATOPlex V3.1 multiplex PCR amplification sequencing. ATOPlex V3.1 adopts a double insurance design with dual primer coverage. Artificially synthesized DNA is spiked in during the multiplex PCR amplification as quality control (external control) for library construction and assisting quantification. Furthermore, the human glyceraldehyde-3-phosphate dehydrogenase gene (GAPDH) is also detected as quality control for sample processing and nucleic acid extraction, and also as an internal reference for relative quantification of viral nucleic acid in each sample (Fig.1).

**Fig 1.**
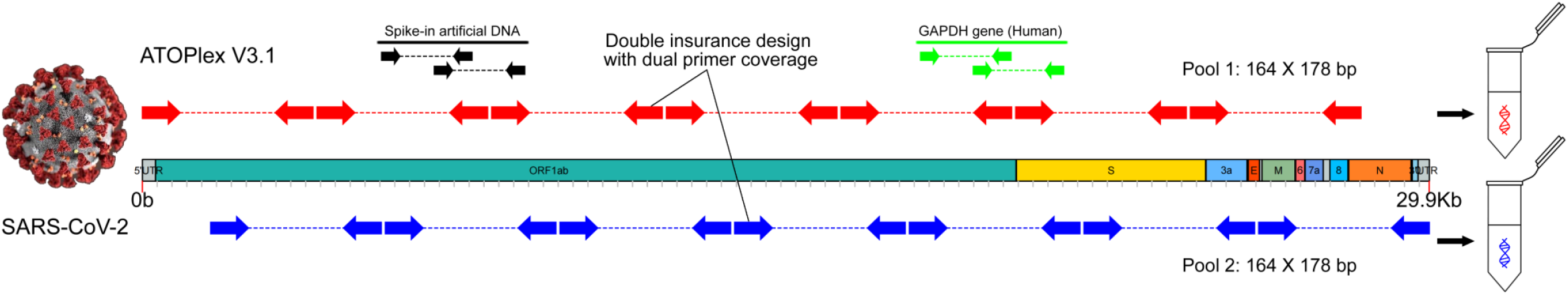
Mechanism of ATOPlex sequencing. ATOPlex is a multiplex PCR amplicon-based high-throughput sequencing platform, which can detect the whole genome of trace microorganisms (especially viruses with smaller genomes) in complex samples. ATOPlex V3.1 adopts a double insurance design with dual primer coverage. Artificially synthesized DNA is spiked in during the multiplex PCR amplification as quality control for library construction and assisting quantification. Furthermore, the human glyceraldehyde-3-phosphate dehydrogenase gene (GAPDH) is also detected as quality control for sample processing and nucleic acid extraction, and also as an internal reference for relative quantification of viral nucleic acid in each sample.

The RNA multiplex PCR amplicon libraries were constructed by ATOPlex SARS-CoV-2 Full Length Genome Panel following the manufacture’s protocol (MGI, Shenzhen, China). Briefly, sewage RNA extract (10 μL) was converted to the first-strand cDNA by reverse transcriptase with random hexamers (5’-NNNNNN-3’). The reverse transcription was performed in a Veriti 96 Well Thermocycler (Applied Biosystems, USA) using the following procedure: 5 min at 25°C, 20 min at 42°C, 5 min at 95°C. The reverse transcript of each sample was mixed with a fixed copy number of artificially synthesized lambda genomic DNA (external control). Thereafter, the mixture was bisected into two equal parts (10 μL each) and two parallel PCR amplifications were performed using two independent primer pools (PCR Primer Pool 1 V3.1 and PCR Primer Pool 2 V3.1), respectively. The PCR primers in two primer pools include (1) primers dual-covering the whole genome of SARS-CoV-2, (2) primers targeting the spiked synthesized lambda DNA (external control) and (3) primers targeting the human GAPDH gene (internal control). The primer sequences (V2.0) are available at on online https://github.com/MGI-tech-bioinformatics/SARS-CoV-2_Multi-PCR_v1.0/blob/master/database/nCoV.primer.2.0.xls. The PCR amplifications were performed in a Veriti 96 Well Thermocycler (Applied Biosystems, USA) using the following procedure: 5 min at 37°C, 5 min at 95°C, 35 amplification cycles (consisting of 10 s at 95°C, 1 min at 62°C, 1 min at 58°C and then 20 s at 72°C), followed by a final elongation step at 72 °C for 1 min. The amplification products from two primer pools were mixed and purified with MGIEasy DNA Clean Beads. The amplification products were fragmentized, end-polished, A-tailed, and ligated with the adaptors.

PCR products of samples were pooled at equimolar and converted to single-stranded circular DNA (ssDNA) with the MGIEasy Circularization Kit (MGI, China). These circularized DNA went through rolling circle amplification to form billions of DNA nanoballs (DNBs). DNBs-based libraries were sequenced on DNBSQE-E5 platform with paired-end 100 nt strategy.

### 2.5. Sequencing data analysis

After ATOPlex multiplex PCR amplicon sequencing, MGI MetargetCOVID VX.X software is used for data analysis. The internal process was that low quality reads were filtered and then the clean reads were mapped against the SARS-CoV-2 reference genome (WuHan-Hu-1, NC_045512.2). The reads of mapped files contained primer will be trimmed by using an in-housed script. Then the SNPs (single-nucleotide polymorphisms), indels (insertions and deletions), MNPs (multi-nucleotide polymorphisms), and complex events (composite insertion and substitution events) were called based on the BAM file. After that, a consensus genome was obtained in term of the variant calling VCF file and the SARS-CoV-2 reference genome. Finally, the clades were provided through mutation identification, consensus genome quality checking, and phylogenetic placement and visualization to assign the lineages.

The sequencing data gave the reads of spike-in external control (artificially synthesized lambda genomic DNA), internal reference (GAPDH) and SARS-CoV-2. As the copy number of spike-in external control was known, the concentration of SARS-CoV-2 nucleic acid in each RNA extracts was calculated accordingly.

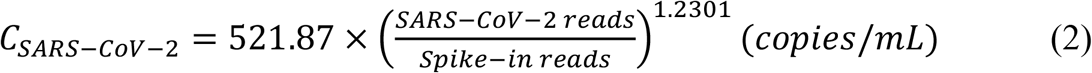

Parameters 521.87 and 1.2301 were provided by the sequencing company derived from previous systematic experimental study. The relative abundance of SARS-CoV-2 nucleic acid against the internal reference (GAPDH) was calculated with the following equation.

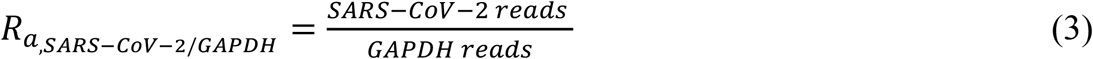

### 2.6. Deconvolution of virus variants for sewage samples

To ensure the reliability of the results, two independent methods were used to deconvolute sewage sequencing data to resolve multiple SARS-CoV-2 lineages in sewage samples. The sequencing company MGI used Freyja for the deconvolution, which is based on a depth-weighted de-mixing method [11]. The relative abundance of SARS-CoV-2 variants/lineages in sewage samples was provided to us as an Excel spreadsheet. The second method was developed in in this work, which was amended from the first one. The deconvolution process was illustrated in Supplementary Fig. S1. The mutation sites with probability more than 1% were recorded when mapping the sequencing reads against the reference sequence, and those sites with sequencing depth low than 100 were filtered. The filtered mutation probability of each site was recorded in a vector ***P***.

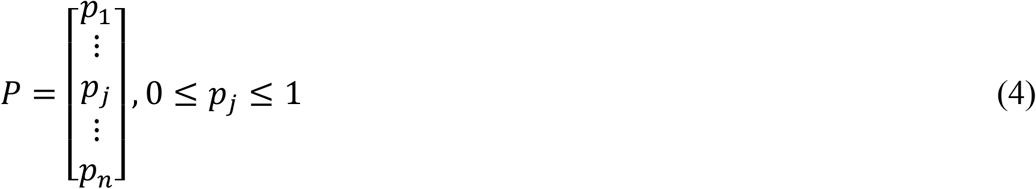

where ***p***_***j***_ represents the overall mutation probability of site ***j***, which is a linear combination of mutation of each virus lineage at the mutation site. The “Barcode” matrix ***A*** containing lineage-defining mutations for known virus lineages was still in use.

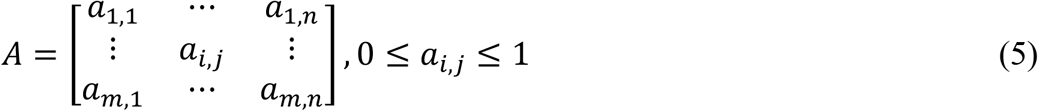

The matrix was obtained from the UShER global phylogenetic tree using the matUtils package [32], where the elements ***a***_***i***,***j***_ denotes the mutation of virus lineage ***i*** at site ***j***. Since each virus lineage is defined by a combination of specific mutation sites, each row of the matrix represents one lineage (out of more than 2,400 lineages included in the UShER global phylogenetic tree), and the distribution of individual mutations among different lineages is represented as columns. This matrix was also filtered to exclude those mutation sites (columns) and lineages (rows) undetected in all sewage samples. The relative abundance of virus lineages in sewage is specified with a vector ***X***.

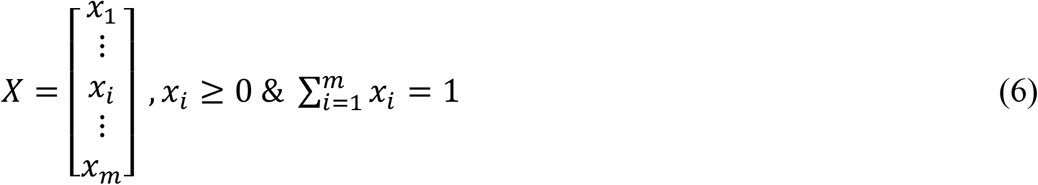

Where ***x***_***i***_ represents the relative abundance of virus lineage ***i***. The relative abundance vector ***X*** for each sewage sample can be obtained by solving the following constrained minimum problem.

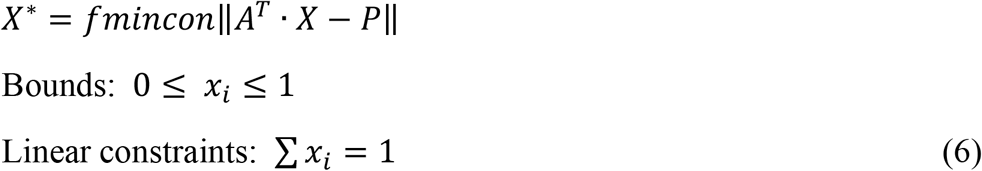

The problem was solved using the built-in functions in MATLAB R2017a (MathWorks, United States). Please refer to the Supplementary MATLAB script “V_POWER.m” and the attached dataset “Mutation.xlsx”.

### 2.7. Data availability

The sequencing data (clean FastQ reads) reported in this paper have been deposited in the Genome Sequence Archive [33] in National Genomics Data Center [34], China National Center for Bioinformation (CNCB) / Beijing Institute of Genomics (BIG), Chinese Academy of Sciences (GSA: CRA011084 or BioProject: PRJCA017073) that are publicly accessible at https://ngdc.cncb.ac.cn/gsa.

## 3. Results

### 3.1. Relative abundance of SARS-CoV-2 nucleic acid in sewage samples

When quantifying SARS-CoV-2 nucleic acid in the sewage samples, the abundance of an endogenous reference biomarker crAssphage was co-quantified. The relative abundance of SARS-CoV-2 nucleic acid was calculated to be around 10^−5^-10^−4^ GC/crAssphage GC (Fig. 2.). The concentration of crAssphage in sewage was reported to be about 10^9^ GC/L [35]. Thus, the concentration of SASR-CoV-2 RNA is about 10^4^- 10^5^ GC/L. There seems to be no close linear correlation between the abundance of the two target genes (ORF1ab and N genes) of SARS-CoV-2 in sewage. The relative abundance of SARS-CoV-2 against the human GAPDH gene was calculated according to the ATOPlex sequencing reads via Eq. (2). The calculated relative abundance of SARS-CoV-2 nucleic acid (***R***_***a, SARS-CoV-2/GAPDH***_) in the sewage experienced a process of increasing first and then decreasing (insect of Fig. 2A).

**Fig 2.**
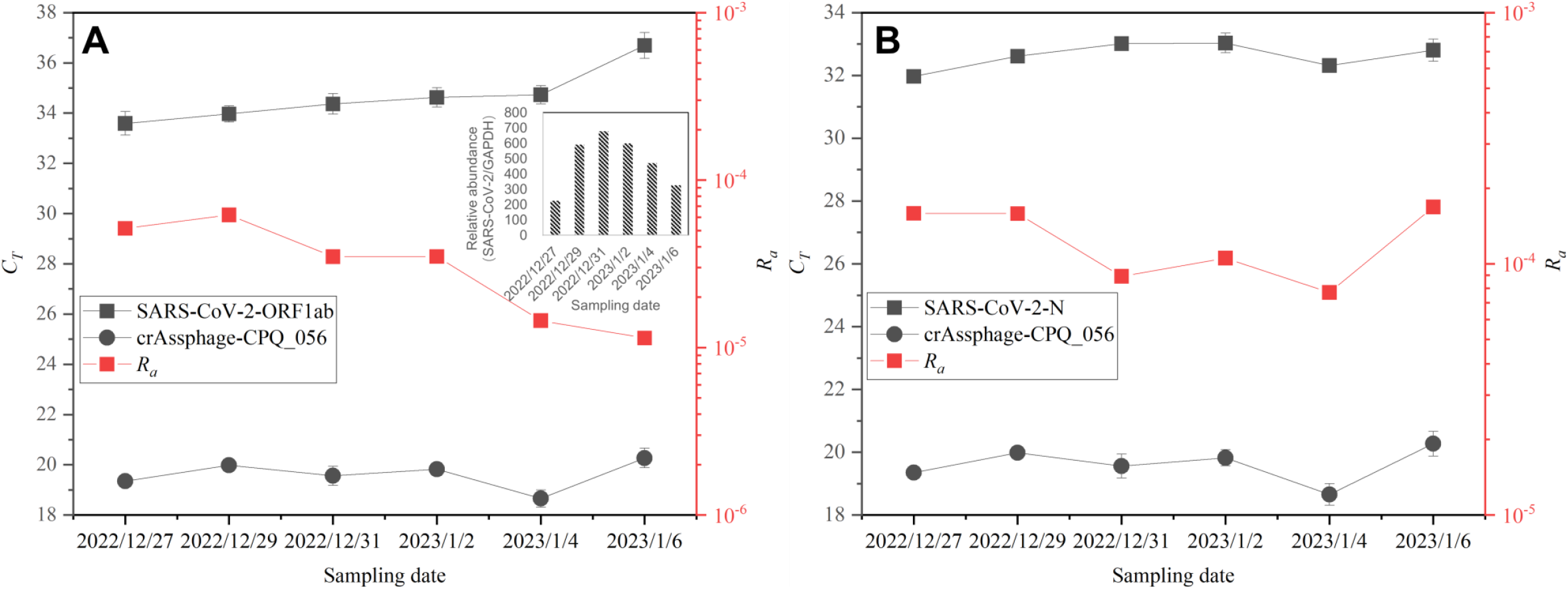
Relative abundance of SARS-CoV-2 genes in sewage samples. The qPCR CT values of SARS-CoV-2 genes and the endogenous biomarker crAssphage are exhibited. The calculated relative abundances (***R***_***a***_) of ORF1ab and N gene are shown in panel A and B, respectively. The inset in panel A gives the relative abundance of SARS-CoV-2 nucleic acids calculated based on ATOPlex sequencing results. The internal reference for relative quantification is the human glyceraldehyde-3-phosphate dehydrogenase gene (GAPDH).

### 3.2. Multiplex PCR amplicon sequencing reflects variant prevalence in sewage

ATOPlex multiplex PCR amplicon sequencing gave good genome coverage and high sequencing depth for all six sewage samples (Fig. 3). Most of the sequencing reads (≥90%) were from SARS-CoV-2 genome (Table 2). Except for 3’-UTR and 5’-UTR, the average sequencing depth of most other genome regions is more than 1000 x (Supplementary dataset 1). The 100 x coverages for six sewage samples are overall greater than 98.5% (Table 2). Except for two samples (2023/1/4 and 2023/1/6) with amplicon drop off in the E gene (the last two panels in Fig. 3), the 100 x coverage of the SARS-CoV-2 genome coding region exceeded 99% (Supplementary dataset 1). We also found that the sequencing coverage/depth plots of different sewage sample exhibited similar “waveform” (Fig. 3). This reflects the ability of the two primer pools to cover the entire genome sequence of SARS-CoV-2 and the bias of PCR amplification. Overall, the coverage of the viral genome by the multiplex PCR is good.

**Table 2.** Basic information of ATOPlex sequencing results.

| Sample ID | GAPDH reads | Spike-in control reads | SARS-CoV-2 reads | SARS-CoV-2 (copies/mL) | Relative abundance SARS-CoV-2/GAPDH | 100 x Coverage (%) | Average depth | Total mutation | Lineage ID |
| --- | --- | --- | --- | --- | --- | --- | --- | --- | --- |
| 2022/12/27 | 3514 | 72585 | 791966 | 9868.1 | 225.37 | 99.17% | 2124.08 | 77 | BA.5.2 |
| 2022/12/29 | 1467 | 95434 | 867871 | 7887.21 | 591.60 | 99.42% | 2303.02 | 80 | BA.5.2 |
| 2022/12/31 | 1029 | 54149 | 697512 | 12103.91 | 677.85 | 99.25% | 1888.88 | 80 | BA.5.2 |
| 2023/1/2 | 940 | 44298 | 563937 | 11929.82 | 599.93 | 98.81% | 1510.35 | 80 | BA.5.2 |
| 2023/1/4 | 1032 | 56414 | 486036 | 7379.93 | 470.97 | 98.60% | 1288.6 | 77 | BA.5.2 |
| 2023/1/6 | 3706 | 108615 | 1204748 | 10069.89 | 325.08 | 99.09% | 3166.26 | 77 | BA.5.2 |

GAPDH: The internal reference gene encoding human glyceraldehyde-3-phosphate dehydrogenase.

**Fig 3.**
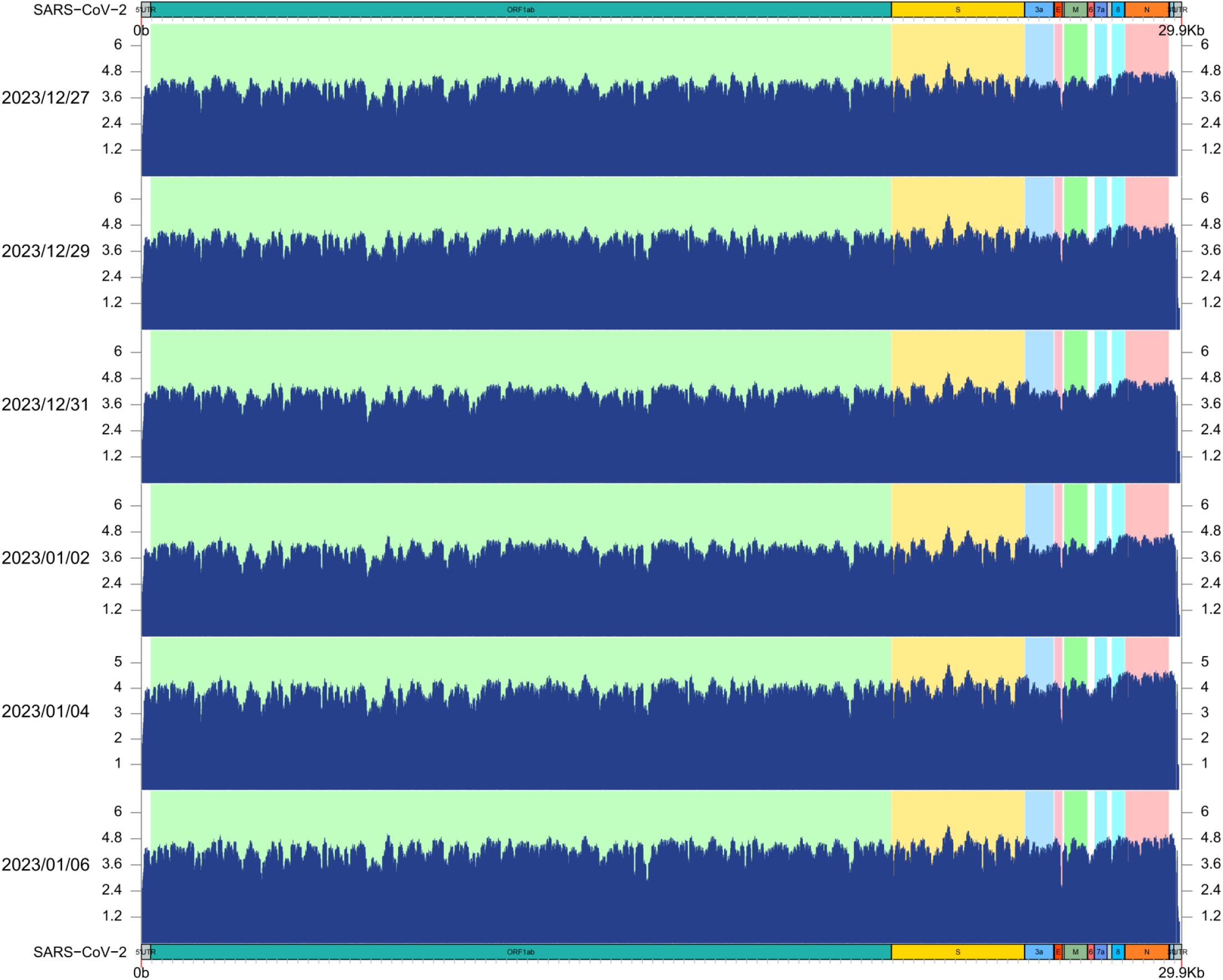
SARS-CoV-2 genome coverage and sequencing depth of ATOPlex multiplex PCR amplicon sequencing for six sewage samples. The horizontal coordinate is the nucleic acid sequence position of the viral genome, and the vertical coordinate is the logarithm value of the sequencing depth (log_10_). The genome coverage is good and the sequencing depth is high. Except for 3’-UTR and 5’-UTR, the average sequencing depth of other regions is more than 1000 ×.

Sample deconvolution gives the relative abundance of SARS-CoV-2 lineages in sewage samples. Two deconvolution methods give similar results (Fig. 4A and B). The main virus variants (prevalence rate>1%) in sewage samples exhibit clear evolutionary genetic correlations (Fig. 4C). They all evolved from the BA. 5.2 lineage through a series of mutations. During the process of evolution, three main sublineages were formed, namely BA.5.2.49, BA.5.2.48, and BF.7.14. Further evolution of each sublineage has led to the emergence of new variants. The results obtained by the two deconvolution methods differ at the haplotype level but are highly consistent at the sublineage level (Fig. 4A and B).

**Fig 4.**
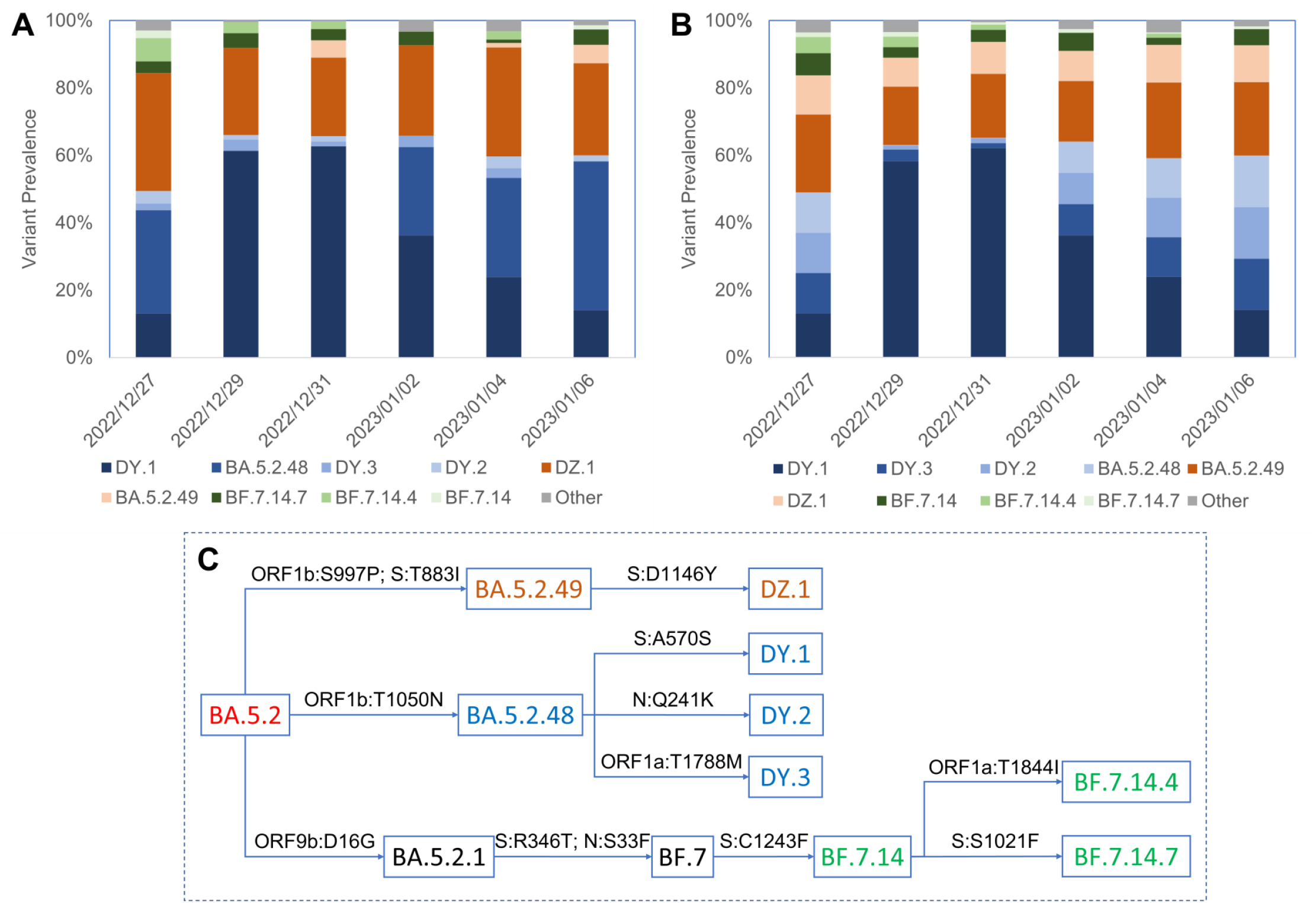
The prevalence (%) of virus variants/lineages in sewage samples. The results obained with Freyja and our method were ehibited in panels **A** and **B**, respectively. The main virus variants (Prevalence%>1%) in sewage samples exhibit clear evolutionary genetic correlations. They all evolved from the BA. 5.2 lineage through a series of mutations. During the process of evolution, three main sublineages were formed, namely BA.5.2.49, BA.5.2.48, and BF.7.14 (Panel **C**). The virus variants of the same sublineages are indicated with same colors of different saturations in panels A and B. Blue represents the sublineage BA.5.2.48, brown represents BA.5.2.49, and green represents BF.7.14. In addition, the sampling time is very consistent with the timeline of uploading the nucleic acid sequences of the corresponding virus variants/lineages from Sichuan to the database (GISAID, https://gisaid.org/) (Supplementary Table S2). This indicates the reliability of the sewage sequencing results.

### 3.3. Wastewater reveals local community transmission of SARS-CoV-2 variants

To prove the reliability of the sewage sequencing data, we retraced the pango-designation issues of these virus variants/lineages detected in these sewage samples on the website https://cov-lineages.org/lineage_list.html and recorded their information in Supplementary Table S2. The amino acid mutation sites of these virus variants/lineages are exhibited in Supplementary Fig. S2. Only the mutation frequencies of mutation sites in ORF1ad and S gene with relatively concentrated mutation are shown. These mutation sites were all detected in these sewage samples (Supplementary dataset 2). The main transmission sites (“Most common countries”) of these SARS-CoV-2 variants/lineages are all in China. Issue-open dates of these lineages were concentrated on December 26 and 27, which were around our sampling date. We also found that 26 sequences of BA.5.2.49 were ever uploaded from Sichuan on Dec 29^th^, 2022 and two more sequences were uploaded from Sichuan on Jan 8^th^, 2023. The branch of lineage BF.7.14.7 contains 11 out of 34 (33%) Sichuan sequences as of Jan 9th 2023. These facts prove that the sewage viral sequencing data reveals local community transmission of SARS-CoV-2 variants.

## 4.. Discussion

During the multiplex PCR amplification, ***fixed copy number*** of artificially synthesized lambda genomic DNA was spiked in the reaction mixture as ***external control***. The endogenous biomarker human GAPDH gene was also detected as ***internal reference***. The spike-in external control can realize the absolute quantification of SARS-CoV-2 nucleic acid in the sewage RNA extracts. However, RNA extraction may be affected by the nature of the sewage. The concentration of viral nucleic acid in RNA extracts may not reflect the true contents of the virus in sewage samples. The relative abundance of the viral nucleic acid against the endogenous biomarker can reflect the true viral load in wastewater. At the beginning of its development, ATOPlex was mainly used for the detection of clinical samples, so human GAPDH gene was adopted as the internal reference [24]. For sewage samples, commonly used endogenous biomarkers such as human gut specific bacteriophage (crAssphage) [36] and pepper mild mottle virus (PMMoV) [37] may be more suitable as internal references. The RT-qPCR determined abundances of different genes (ORF1ab and N) are not consistent with each other (Fig. 2A and B), and also differ from the abundance determined by ATOPlex sequencing (inset of Fig. 2A). It has been suggested that RT-qPCR may give false negative detection for SARS-CoV-2 [38], which can be attribute to degradation of SARS-CoV-2 RNA in the wastewater matrix [9]. In addition, the accuracy of RT-qPCR quantification is also affected by mutations in the viral genome sequence targeted by primers/probes [39, 40], especially for samples such as sewage that may contain multiple virus variants. Therefore, a quantification method that targets the whole genome of SARS-CoV-2 is valuable for WBE [41]. The ATOPlex sequencing results indicate that the relative abundance of SARS-CoV-2 RNA in wastewater increases first and then decreases over time (inset of Fig. 2A), which seems reasonable.

The design of the primer set is crucial for improving the sequencing coverage and depth of multiplex PCR amplicon sequencing [42]. The amplicon droop-offs due to primer dimers and primer mismatch caused by virus genome mutations are problems that needs to be addressed during primer set design and optimization [42]. To improve the coverage of sequencing, the primer set is designed to tile amplicons across entire sequence of the published reference SARS-CoV-2 genome NC_045512.2 [43]. The primer set is divided into two separate subsets (Pools 1 and 2), and primer pairs targeting adjacent regions are separated in alternate pools so that overlap of PCR fragments occurs between pools but not within [25, 44]. The ATOPlex SARS-CoV-2 full-length genome panel has evolved to the third version. Its previous versions have shown considerable application potential in the detection of input virus variants [45], quantification of sewage SARS-CoV-2 RNA [41], and resolution of sewage virus lineages [46, 47]. ATOPlex V3.1 also adopts a double insurance design with dual primer coverage (Fig. 1). Analyzing the prevalence of virus lineages in wastewater places higher demands on sequencing coverage and depth. To accurately analyze virus lineages with relative prevalence exceeding 1%, the sequencing depth should reach at least 1000 x. The sequencing coverage and depth of each sewage sample in this study were good with little coverage bias (Supplementary dataset 1), basically meeting the above requirements. The DNB-based sequencing method used in this study, which obtained the sequencing library through rolling circle amplification, ensured the accuracy of sequencing to a considerable extent. Considering that the coverage and depth of sequencing are mainly determined by the design of primer sets, during the deconvolution process, only mutation points with sequencing depths below 100 were filtered, without using the depth-weighted method [11].

Six sewage samples contain three major SARS-CoV-2 variants, while BA.5.2.48 is absent from the samples of 2022/12/29 and 2022/12/31. We obtained sewage samples from a manhole in the student dormitory area. The community monitored by the sewage manhole is very limited, only several student dormitory buildings. Therefore, the secretions of infected individuals contained in these sewage samples have a significant randomness. There may be a situation where the sewage sample collected on a certain day lacks the secretion of an infected person. However, even so, only six sewage samples may already contain important information about the main prevalent SARS-CoV-2 lineages in the entire region (Chengdu) at that time.

Multiplex PCR amplicon sequencing has both quantitative and qualitative potential and is currently the most powerful WBE tool. Overall, our study is still a retrospective one. The premise for defining a SARS-CoV-2 lineage is that the virus variant should have the potential of ongoing transmission, which means it has achieved a certain scale of community transmission when being named [48]. When new SARS-CoV-2 variants were detected in sewage, the corresponding lineages had not yet been defined. As in the case of this study, the sampling time for sewage spans from Dec. 27, 2022 to Jan. 6, 2023, and the main SARS-CoV-2 lineages detected in sewage have not yet been defined during that period. Multiple PCR amplicon sequencing should enable the real-time tracking of the transmission of known virus lineages imported into a new community. In addition, there is still room for optimization in the deconvolution method of sewage samples. Different sequencing methods may require targeted deconvolution methods. However, it is worth noting that any deconvolution method cannot distinguish recombination events between virus variants. Therefore, in summary, clinical testing and sewage testing are complementary in community monitoring of infectious diseases. WBE cannot completely replace clinical testing, but in terms of large-scale monitoring, WBE has shown unique advantages over clinical testing. In the long run, population growth and global warming are likely to exacerbate human–pathogen interactions, combined with the continuous evolution of pathogens, suggesting an urgent need for continuous technological advancements to keep up with these changes and provide more effective surveillance. When the next new threat arises, a universal early-warning system that includes sewage pathogen variant monitoring is crucial for protecting the public and relieving the pressure on healthcare system [49].

## 5. Conclusion

- During the outbreak of infection in Chinese Mainland at the end of 2022, six sewage samples collected from the student dormitory area on Wangjiang Campus of Sichuan University were positive for SARS-CoV-2 nucleic acid via RT-qPCR detection. The abundance of the viral nucleic acid in these sewage samples is approximately 10^4^-10^5^ GC/L.
- ATOPlex multiplex PCR amplicon sequencing depicted the community circulation and evolution of SARS-CoV-2 variants.
- Multiplex PCR amplicon sequencing is up to date the most powerful WBE tool, which can quantitatively reveal the community transmission of virus variants.

## Data Availability

All data produced in the present study are available upon reasonable request to the authors

## CRediT authorship contribution statement

**Langjun Tang:** Methodology, Investigation, Data curation, Visualization, Writing - review & editing. **Zhenyu Guo:** Data curation, Visualization, Writing - review & editing. **Xiaoyi Lu**: Writing - review & editing. **Junqiao Zhao**: Writing - review & editing. **Yonghong Li:** Conceptualization, Supervision, Project administration, Writing - review & editing. **Kun Yang:** Conceptualization, Funding acquisition, Supervision, Project administration, Methodology, Visualization, Writing - original draft, Writing - review & editing.

## Acknowledgements

We thank the financial supports from the National Natural Science Foundation of China (No. 22176133, No. 21677104). We would like to express our gratitude to Mr. Bo Zhang at MGI for his assistance in sequencing data processing.

## Appendix A. Supplementary information

Doc. 1. Protocol for qPCR.

Doc. 2. Protocol for RT-qPCR.

Supplementary Fig. S1. Diagram illustrating the deconvolution process of sewage ATOPlex sequencing data in this work.

Supplementary Fig. S2. The mutation sites of main virus variants/lineages detected in the sewage samples recorded in the database. Only the mutation sites in ORF1ad and S gene are shown. The data was obtained from China National Center for Bioinformation (CNCB) at https://ngdc.cncb.ac.cn/ncov/knowledge/compare on May 12th 2023.

Supplementary dataset 1: ATOPlex sequencing detailed genome coverage & sequencing depth information.

Supplementary dataset 2:ATOPlex sequencing detailed mutation information-

Zhenyu Guo needs to update the data.

Supplementary dataset 3: Mutations.xlsx contain data for the deconvolution. Supplementary Table S1. Metadata of ATOPlex sequencing.

Supplementary Table S2. Lineage information from cov-lineage.org.

Supplementary MATLAB script: V_POWER.m for solving the deconvolution.

## Nomenclature

A: Barcode matrix containing lineage-defining mutations for known SARS-CoV-2 lineages
*a*_*i,j*_: the mutation of virus lineage ***i*** at site ***j***
*R*_*a*_*-*: relative abundance of SARS-CoV-2 gene in sewage (against crAssphage CPQ_056)
*R*_*a,SARS-CoV-2/GAPDH*_: relative abundance of SARS-CoV-2 genome in sewage (against GAPDH)
*C*_*SARS-CoV-2*_: concentration of SARS-CoV-2 genome in sewage nucleic acid extracts
*C*_*T*_: cycle threshold
*c*_*0*_: initial copy number of the target gene during qPCR amplification (GC/reaction)
*P*: vectors recording the probability of each mutation site
*p*_*j*_: the overall probability of mutation site ***j***
*t*: hydraulic retention time (HRT) of virus/biomarker in sewage (h)
*t*_*1/2*_: half-life of virus in the sewage (h)
*X*: the vector indicating relative abundance of virus lineages
*X\**: the optimized relative abundance vector *x*_*i*_ the relative abundance of virus lineage ***i***

## Greek letters

*ξ* apparent amplification coefficient, equaling 2 under ideal condition

## Subscripts

*i* virus lineage ***i***

*j* mutation site ***j***

*s* parameters in related to the reference biomarker

*t* parameters in related to the target viral gene

