## Supplementary materials for "Multiplex PCR amplicon sequencing revealed community transmission of SARS-CoV-2 lineages on the campus of Sichuan University during the outbreak of infection in Chinese Mainland at the end of 2022"

### Supplementary Information Table of Contents

#### Supplementary documents

**Doc. 1.** Protocol for qPCR. 3

**Doc. 2.** Protocol for RT-qPCR. 4-5

#### Supplementary figures

**Supplementary Fig. S1.** Diagram illustrating the deconvolution process of sewage ATOPlex sequencing data in this work. 6

**Supplementary Fig. S2.** The mutation sites of main virus variants/lineages detected in the sewage samples recorded in the database. 7

#### Supplementary datasets and tables

1. **Supplementary dataset 1**: ATOPlex sequencing detailed genome coverage & sequencing depth information.
2. **Supplementary dataset 2**: ATOPlex sequencing detailed mutation information. Zhenyu Guo needs to update the data.
3. **Supplementary dataset 3**: “**Mutations.xlsx**” contain data for the deconvolution.
4. **Supplementary Table S1.** Metadata of ATOPlex sequencing.
5. **Supplementary Table S2.** Lineage information from cov-lineage.org. 8

#### Supplementary MATLAB script

**V_POWER.m** for solving the deconvolution is attached in supplementary materials. 8

25

### Supplementary documents

#### 1. Protocol for qPCR

##### *1.1. Kit*

The qPCR kit PerfectStart® Green qPCR SuperMix (Product No. AQ101, TransGen Biotech Co., Ltd, Beijing, China) was used to quantify the reference biomarker crAssphage.

##### *1.2. Composition of reaction mixture*

Reaction mixture for qPCR quantification of the reference biomarker crAssphage.

| Component | Volume (µL) | Final Concentration |
|---|---|---|
| Template | 2 | as required |
| Forward Primer (10 µM) | 0.4 | 0.2 µM |
| Reverse Primer (10 µM) | 0.4 | 0.2 µM |
| 2 X PerfectStart® Green qPCR SuperMix | 10 | 1 X |
| BSA (20 mg/mL) | 0.4 | 0.4 mg/mL |
| Nuclease-free Water | 6.8 | - |
| Total volume | 20 µL | - |

##### *1.3. Thermal cycle conditions*

Thermal cycle setting for qPCR quantification of the reference biomarker crAssphage.

| Steps | Temperature (°C) | Duration | Cycles |
|---|---|---|---|
| Pre-denaturation | 95 | 5 min | 1 |
| Denaturation | 95 | 15 s | 42 |
| Annealing/Extension | 62 | 30 s | |
| Melting curve analysis | According to instrument guidelines | | |

#### 2. Protocol for RT-qPCR

##### *2.1. Kits*

The RT-qPCR kit TransScript® II Probe One-Step qRT-PCR SuperMix (Product

No. AQ321, TransGen Biotech Co., Ltd, Beijing, China) was used to quantify ORF1ab and N of SARS-CoV-2.

##### *2.2. Composition of reaction mixture*

Reaction mixture for RT-qPCR quantification of SARS-CoV-2 ORF1ab and N.

| Component | Volume (µL) | Final Concentration |
| --- | --- | --- |
| Template | 2 | as required |
| Forward Primer (10 µM) | 0.4 | 0.2 µM |
| Reverse Primer (10 µM) | 0.4 | 0.2 µM |
| **Probe (10 µM)** | 1 | 0.5 µM |
| 2 X PerfectStart™ Probe One-Step qPCR SuperMix | 10 | 1 X |
| TransScript® II Probe One-Step RT/RI Enzyme Mix | 0.4 | - |
| BSA (20 mg/mL) | 0.4 | 0.4 mg/mL |
| RNase-free Water | 5.4 | - |
| Total volume | 20 µL | - |

##### *2.3. Thermal cycle conditions*

Thermal cycle setting for RT-qPCR quantification of SARS-CoV-2 ORF1ab and N.

| Steps | Temperature (°C) | Duration | Cycles |
| --- | --- | --- | --- |
| Reverse transcription | 50 | 5 min | 1 |
| Pre-denaturation | 95 | 5 min | 1 |
| Denaturation | 95 | 15 s | 42 |
| Annealing/Extension | 58 | 30 s | |
| Melting curve analysis | **Probe protocol without melting curve analysis** | | |

- **Note:** These qPCR and RT-qPCR were all operated on QuantStudio™ 1 Real-Time PCR System (ThermoFisher Scientific, USA).

**Supplementary Fig. S1.** Diagram illustrating the deconvolution process of sewage ATOPlex sequencing data in this work.

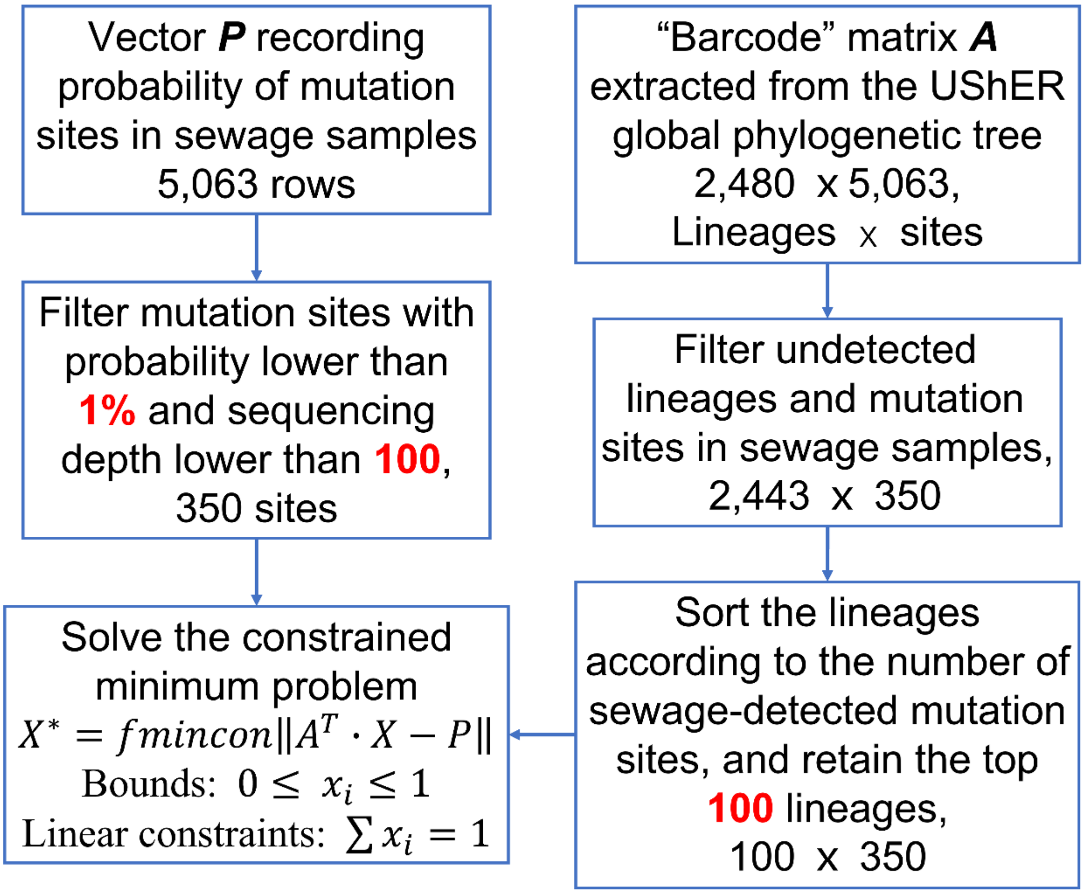

**Supplementary Fig. S2.** The mutation sites of main virus variants/lineages detected in the sewage samples recorded in the database. Only the mutation sites in ORF1ad and S gene are shown. The data was obtained from China National Center for Bioinformation (CNCB) at [https://ngdc.cncb.ac.cn/ncov/knowledge/compare](https://ngdc.cncb.ac.cn/ncov/knowledge/compare) on May 12th 2023.

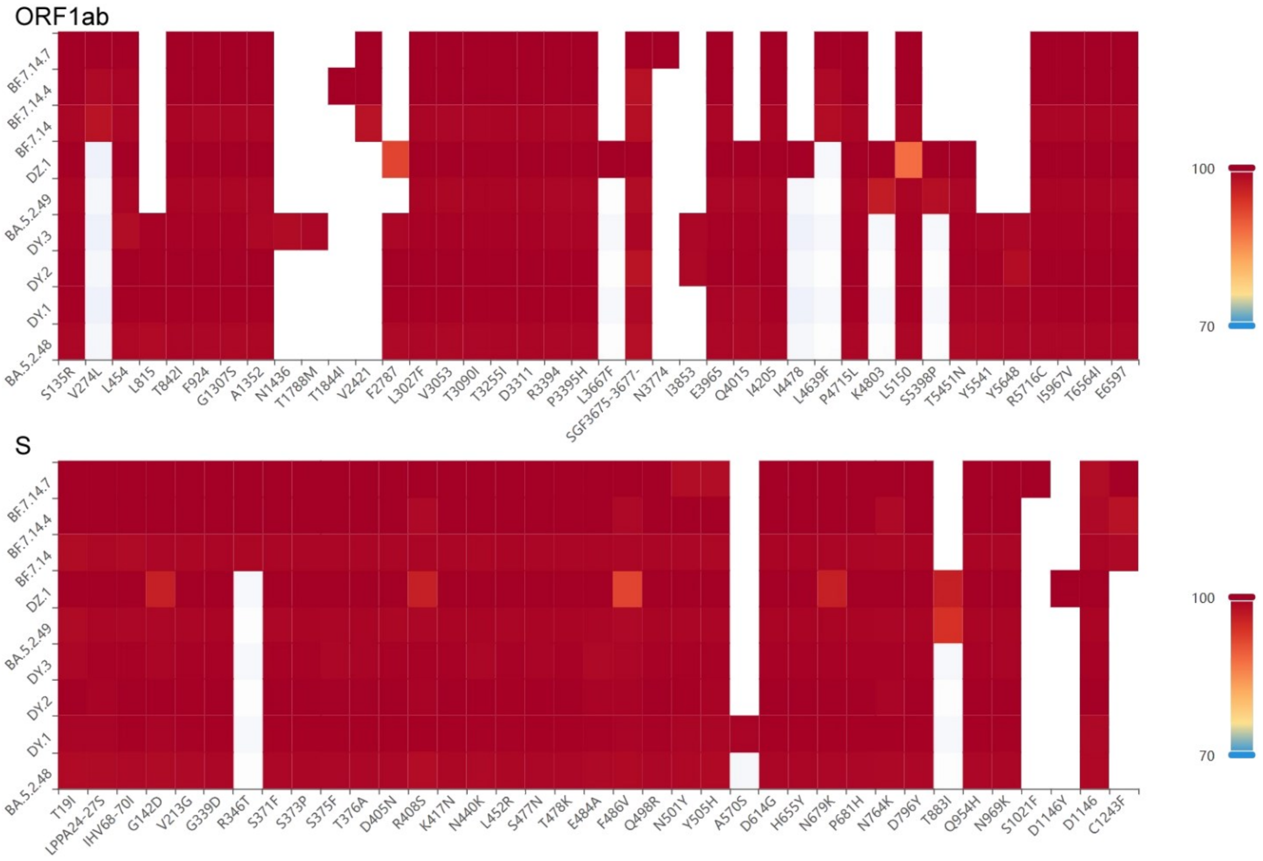

#### Supplementary datasets and tables

1. **Supplementary dataset 1**: ATOPlex sequencing detailed genome coverage & sequencing depth information.
2. **Supplementary dataset 2**：ATOPlex sequencing detailed mutation information. Zhenyu Guo needs to update the data.
3. **Supplementary dataset 3**: “**Mutations.xlsx**” contain data for the deconvolution.
4. **Supplementary Table S1.** Metadata of ATOPlex sequencing.
5. **Supplementary Table S2.** Lineage information from cov-lineage.org.

**Note:** All supplementary datasets and tables are provided in separate excel spreadsheets.

#### Supplementary MATLAB script

**“V_POWER.m”** for solving the deconvolution is attached in supplementary materials, which is used in conjunction with the Excel spreadsheet “**Mutations.xlsx”.**
